# Shielding under endemic SARS-CoV-2 conditions is easier said than done: a model-based analysis

**DOI:** 10.1101/2023.01.22.23284884

**Authors:** Madison Stoddard, Lin Yuan, Sharanya Sarkar, Matthew Mazewski, Debra van Egeren, Shruthi Mangalaganesh, Ryan P. Nolan, Michael S. Rogers, Greg Hather, Laura F. White, Arijit Chakravarty

**Affiliations:** Fractal Therapeutics, Cambridge, MA, USA; Dartmouth College, Hanover, NH, USA; Independent Researcher; Stanford University School of Medicine, Stanford, CA, USA; Monash University, Melbourne, VIC, AUS; Halozyme Therapeutics, San Diego, CA, USA; Harvard Medical School, Boston, MA, USA; Boston Children’s Hospital, Boston, MA, USA; Sage Therapeutics, Cambridge, MA, USA; Boston University School of Public Health, Boston, MA, USA

## Abstract

As the COVID-19 pandemic continues unabated, many governments and public-health bodies worldwide have ceased to implement concerted measures for limiting viral spread, placing the onus instead on the individual. In this paper, we examine the feasibility of this proposition using an agent-based model to simulate the impact of individual shielding behaviors on reinfection frequency. We derive estimates of heterogeneity in immune protection from a population pharmacokinetic (pop PK) model of antibody kinetics following infection and variation in contact rate based on published estimates. Our results suggest that individuals seeking to opt out of adverse outcomes upon SARS-CoV-2 infection will find it challenging to do so, as large reductions in contact rate are required to reduce the risk of infection. Our findings suggest the importance of a multilayered strategy for those seeking to reduce the risk of infection. This work also suggests the importance of public health interventions such as universal masking in essential venues and air quality standards to ensure individual freedom of choice regarding COVID-19.

## Introduction

The emergence of SARS-CoV-2 has led to high levels of ongoing disease transmission, with repeated waves of viral variants creating an unpredictable trajectory for the ongoing pandemic. At this point, SARS-CoV-2 variants have a very high intrinsic reproductive number (R_0_) ^1^, a strong propensity for asymptomatic spread ^2^ and an ability to evade pre-existing immunity ^1^, making disease suppression challenging. Meanwhile, many countries have scaled back or abandoned universal public health measures designed to slow SARS-CoV-2 spread ^3–6^. Specifically, public health agencies in many countries have removed disease-management measures such as mandatory and/or publicly funded non-pharmaceutical interventions (NPIs: masks, testing, contact tracing, quarantines) ^3^. Concurrently, processes for case reporting have also been degraded, with the availability of rapid antigen testing leading to a larger fraction of positive tests going unreported ^7–10^.

A common framing of the public health position in many countries is that managing COVID-19 infection risk is now a matter of “personal responsibility” ^6,11,12^. However, the practical feasibility of reducing one’s risk from SARS-CoV-2 through personal protective measures remains an open question. When it comes to NPIs, one-way masking is substantially less effective than universal masking ^13,14^. According to the CDC, individuals wearing N95 respirators at all times in public indoor settings benefited from 83% reduction in risk of infection under early pandemic conditions ^15^. Thus, the removal of mask mandates exposes individuals to unmasked crowds in indoor settings where exposure is unavoidable (such as public transit, schools, grocery stores and medical settings), making it difficult for individuals to opt-out of the additional COVID-19 risk. Current CDC guidelines only recommend universal masking under “medium or high COVID-19 Community Levels” ^16^, defined not only by case counts, but also by hospitalization rates and hospital bed occupancy (both indicators that lag the risk of infection by weeks) ^17^. At the same time, other NPIs have also been removed: quarantines, testing and social distancing guidelines have been dropped ^6,18^. Meanwhile, improved ventilation has not yet been implemented at scale in Western countries ^19,20^. These changes have occurred even as steps are being taken or called for to reduce outdoor services ^21–23^, and employers have increasingly enforced in-person work (“return to office”) policies ^24–28^ leading to outbreaks ^29–32^. Taken together, these changes over the course of the pandemic leaves individuals seeking to reduce their risk of infection with limited options in terms of NPIs.

Individuals wishing to mitigate personal COVID-19 risk using biomedical interventions also have limited effective options. At present vaccines have limited efficacy against infection ^33^ and transmission ^34,35^ due to antibody waning ^36–39^ and viral immune evasion ^40–43^ Infection also carries a risk of post-acute outcomes (“long COVID”), which is only modestly reduced by vaccination ^44,45^ and likely to be a significant driver of public health and societal outcomes with COVID-19 ^46–48^. At the same time, rapid viral immune evasion has limited the utility of existing monoclonal antibody prophylactics ^49,50^. Coupled with the lack of antiviral prophylactics for SARS-CoV-2, this situation leaves individuals seeking to reduce their risk of infection with few biomedical interventions. Masking, vaccine boosters and social isolation remain the primary means of “shielding”, and the extent to which individuals applying these approaches can reduce their infection frequency under endemic conditions is unknown.

The practical utility of shielding as a public health strategy thus remains an open question, even as many countries have moved to implement some variation of the personal-responsibility-focused approach. In this paper, we use a model-based approach combining population pharmacokinetics (PK) with agent-based epidemiological simulations to examine the impact of shielding on risk of infection and long COVID in the population as a whole. We use estimates of heterogeneity in immune protection in the population (derived from a population PK model) as inputs into an agent-based model of a scenario where repeated reinfections occur under endemic SARS-CoV-2 transmission. We were specifically interested in using modeling to examine whether an individual responsibility approach offers meaningful protection to individuals seeking to reduce their risk of SARS-CoV-2 infection and long COVID.

## Results

### Neutralizing antibody persistence is heterogeneous

Studies of antibody kinetics in convalescent individuals suggest inter-individual variation in nAb kinetics ^51–53^, which may imply variation in vaccine protection in the population. To determine the distribution of nAb kinetics in the population over time, we fitted a population PK model to a dataset containing nAb titers in SARS-CoV-2 convalescents ^54^. As shown in Figure 1A, the model provides a good representation of the population’s antibody kinetics. Model parameters and standard errors are provided in Table 1. In a mixed-effects model, fixed effects represent population central estimates while random effects measure variability in the population. Standard errors less than 30% for all parameters except ω_kel_ (population standard deviation of nAb elimination rate) indicates a good fit. An in-depth goodness-of-fit analysis is provided in Supplemental Figure S3.

**Figure 1.:**
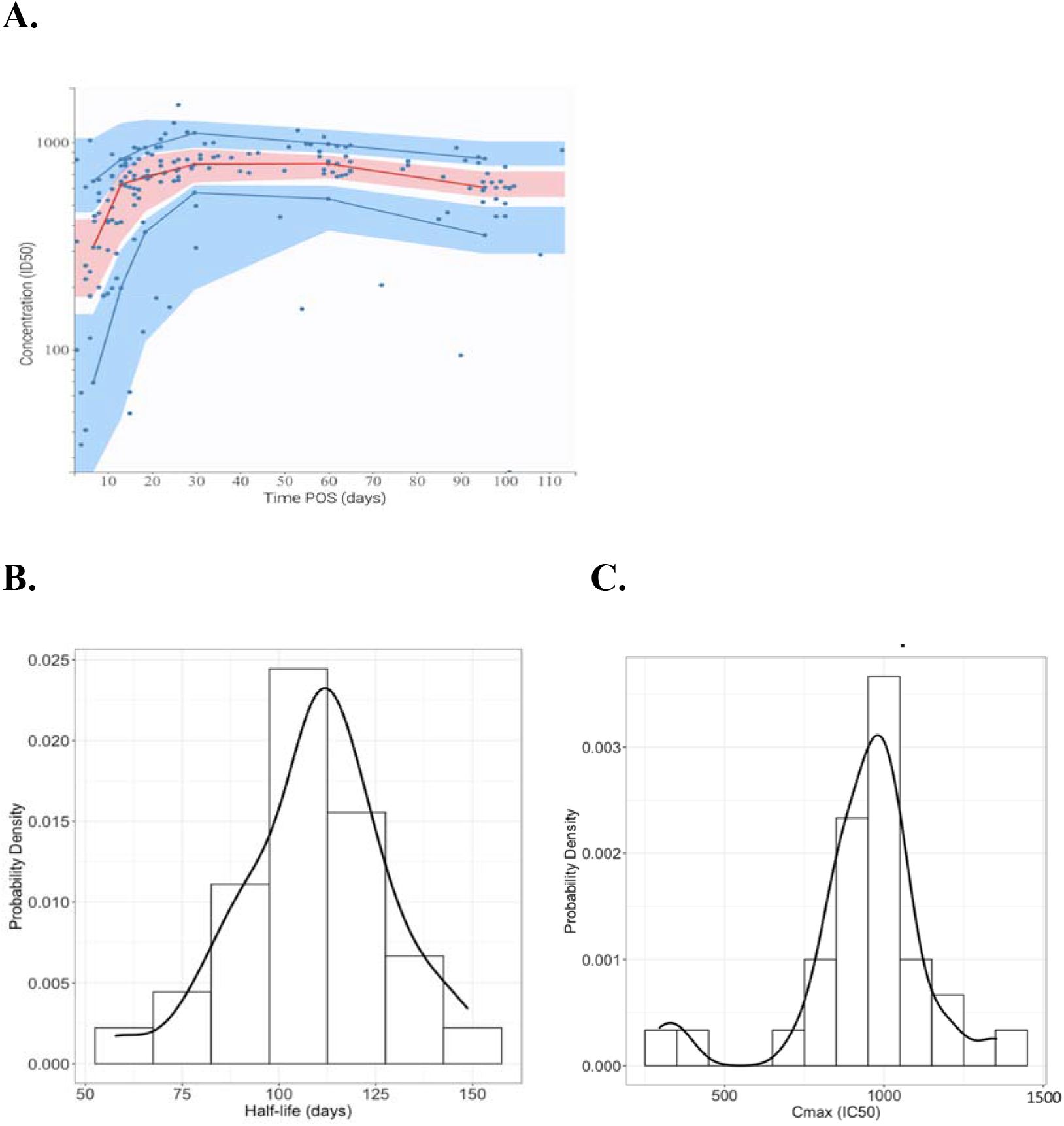
Population PK model fit to anti-SARS-CoV-2 neutralizing antibody titers post-onset of symptoms (POS). **A**. Visual predictive check for model agreement with fitted dataset. Shaded regions represent 90% prediction intervals for model certainty (pink area for the 50^th^ percentile, and blue areas for the 10^th^ and 90^th^ percentiles). Blue dots represent the fitted dataset, with dark blue lines representing the empirical 10^th^ and 90^th^ percentiles and red line representing the empirical 50^th^ percentile. Distribution of **B**. nAb half-life and **C**. nAb peak titer (C_max_) values in the study population.

**Table 1.** Parameter values for fitted nAb kinetics model with standard errors (SE) and relative standard error (RSE).

| Parameter | Value | Units | Standard error | Relative standard error (%) |
| --- | --- | --- | --- | --- |
| *Fixed effects (median)* |  |  |  |  |
| $k_{p, \text{pop}}$ | 38.3 | IC50/days | 5.31 | 13.9 |
| $k_{el, \text{pop}}$ | 0.00628 | 1/days | 0.0012 | 19.1 |
| $T_{in, \text{pop}}$ | 26.9 | days | 26.9 | 15.1 |
| *Standard deviation of the random effects* |  |  |  |  |
| $\omega_k$ | 0.731 | IC50/days | 0.156 | 21.3 |
| $\omega_{kel}$ | 0.346 | 1/days | 0.141 | 40.8 |
| $\omega_{Tin}$ | 0.708 | days | 0.181 | 25.6 |
| *Correlations* |  |  |  |  |
| $\text{corr}_{k, Tin}$ | -0.977 | | 0.0255 | 2.61 |
| *Error model parameters* |  |  |  |  |
| a | 121 |  | 9.11 | 7.51 |

where $\text{corr}_{k, Tin}$ is the correlation between $k_p$ and $T_{in}$ , and a is the coefficient of constant error.

Based on the mixed-effects model fit, the median half-life of neutralizing potency is 109 days (Figure 1B), suggesting that waning will occur within months. Additionally, variability in neutralizing antibody half-life is significant, with a 90% population interval ranging from 79 to 139 days. On a yearly basis, this translates to a range of 6-fold to 24-fold waning (2.6 half-lives per year versus 4.6 half-lives per year). This suggests that neutralizing antibody persistence varies widely in the general population. On the other hand, the median IgG half-life is only 64 days, slightly more than half of the neutralizing potency half-life (Figure S1D). This suggests that affinity maturation may be occurring – that is, antibody potency increases over time after infection. We note that these antibody half-lives are in line with other viral diseases characterized by frequent reinfection, such as common cold coronavirus 229E and influenza (Table S3). This is in contrast to rare reinfection viruses such as measles and rubella, which induce antibodies with half-lives on the order of many years.

The model also showed heterogeneity in peak titer (Figure 1C), with the 90% population interval (90% pi) for peak nAb level spanning from 530 to 1187 multiples of the IC_50_ (nAb titer required to reduce viral infectivity by 50% in a pseudovirus assay).

### Heterogeneity in nAb kinetics results in clinically significant differences in protection from mild and severe COVID-19

In Figure 2, we explore the clinical impact of interindividual heterogeneity in nAb kinetics on protection from strain-matched mild and severe COVID-19. Individual protection from mild and severe COVID-19 is estimated based on nAb titers over time according to the previously established dose-response relationship between nAb titers and COVID-19 protection ^55^. This analysis suggests that protection from COVID-19 is only temporary, even in the absence of immune evasion. For an individual with the median durability of protection, 50% or greater protection from mild COVID-19 persists for less than one year post onset of symptoms (POS) (Figure 2A), while 50% protection from severe disease lasts approximately 22 months (Figure 2B). However, many individuals in the population are predicted to have less durable protection. For example, at the 10th percentile of nAb half-life, protection from mild disease is maintained above 50% for only six months, while greater than 50% severe disease protection lasts just under one year. These outcomes represent the best-case scenario in which loss of protection is driven only by antibody waning; in reality, immune evasion due to viral evolution compounds this rate of immunity loss.

**Figure 2. :**
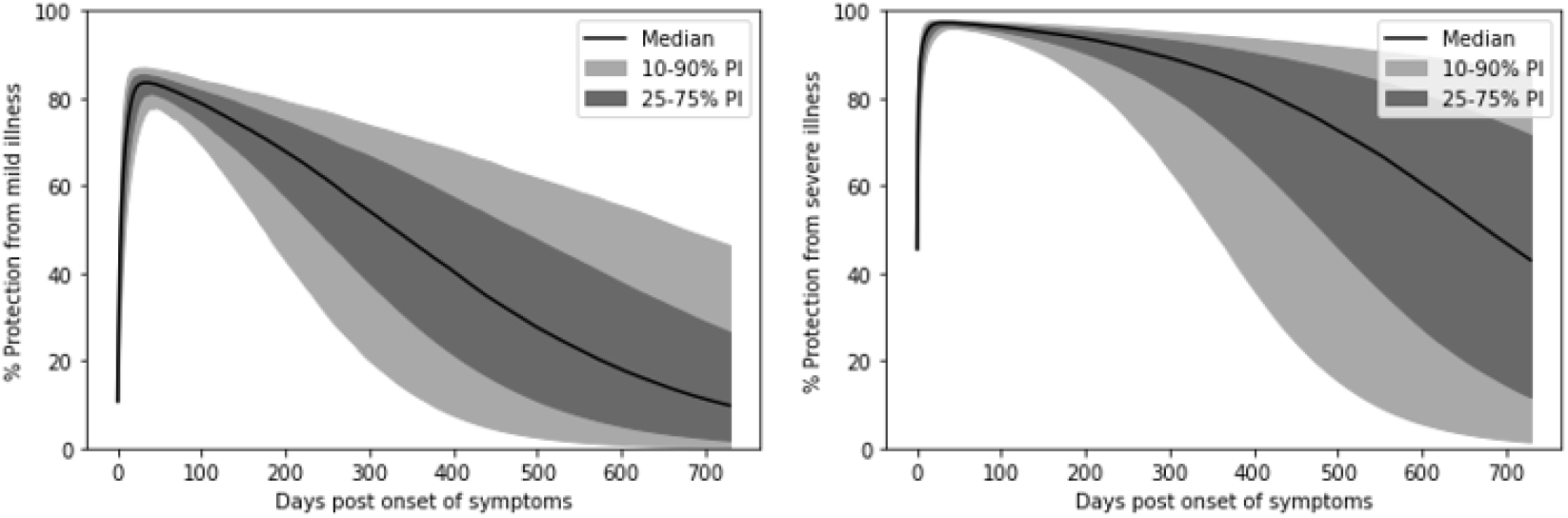
Post-infection protection from mild and severe COVID-19 outcomes. Protection is calculated against strain inducing immunity – that is, immune evasion is not accounted for. Protection over time will be further eroded by immune evasion.

### Under endemic conditions, the range of nAb titers within the population is wide

Using the distribution of nAb half-lives fitted in the mixed effects model, we developed a simplified agent-based model to simulate immunological dynamics under endemic SARS-CoV-2 transmission (Figure 3). This model tracks nAb titers, reinfection frequency, and long COVID risk in a simulated population of 100,000 individuals. These individuals are characterized by varying contact rates, nAb half-lives, and vaccination statuses. For the following analyses, we assumed that 50% of the population is boosted on a once-yearly revaccination schedule. This is consistent with uptake of the first booster shot ^56^, but may be optimistic with respect to future compliance ^57^.

**Figure 3. :**
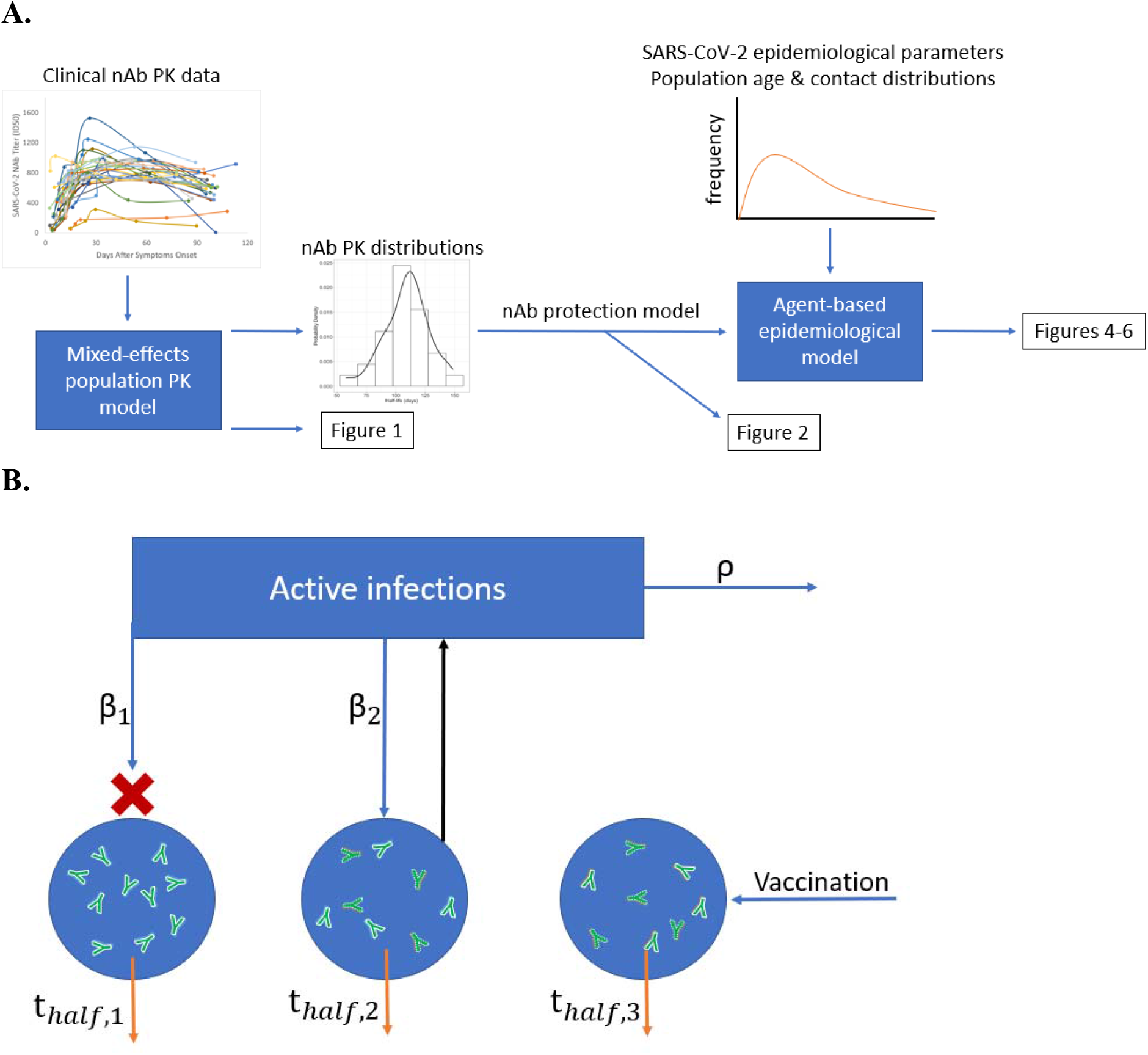
**A**. Information flow diagram demonstrating how models were parameterized and manuscript figures were generated. **B**. Schematic representing the simplified agent-based model. In the model, nAb titer (antibody icons) is tracked for each individual (blue circles) over time. nAb titer decreases over time according to the individual’s nAb half-life and the rate of immune evasion (t_half_), and nAb titer is increased by a fixed multiple on vaccination or infection. Vaccination occurs at a fixed interval, while infection occurs upon successful exposure. The number of exposures over time is determined by the number of active infections and the R_0_. Individuals in the simulated population are stochastically selected for exposure with a probability proportional to their contact rates (β_1_,β_2_). Exposure successfully leads to infection with a probability determined by the individual’s nAb titer upon exposure. If successful, an infection increases the count of active infections by one. Infections resolve according to the rate of recovery ρ.

The agent-based model predicts that under steady-state (endemic) SARS-CoV-2 spread, a wide range of nAb titers are observed in the population, spanning 0.1 convalescent plasma (CP) titer at the 10^th^ percentile to 25 CP titer at the 90^th^ percentile (Figure 4). Additionally, approximately two-thirds of the population are expected to have greater than 1x CP nAb titer at any given time. This is due to the build-up of nAbs across repeated reinfections and vaccinations.

**Figure 4.**
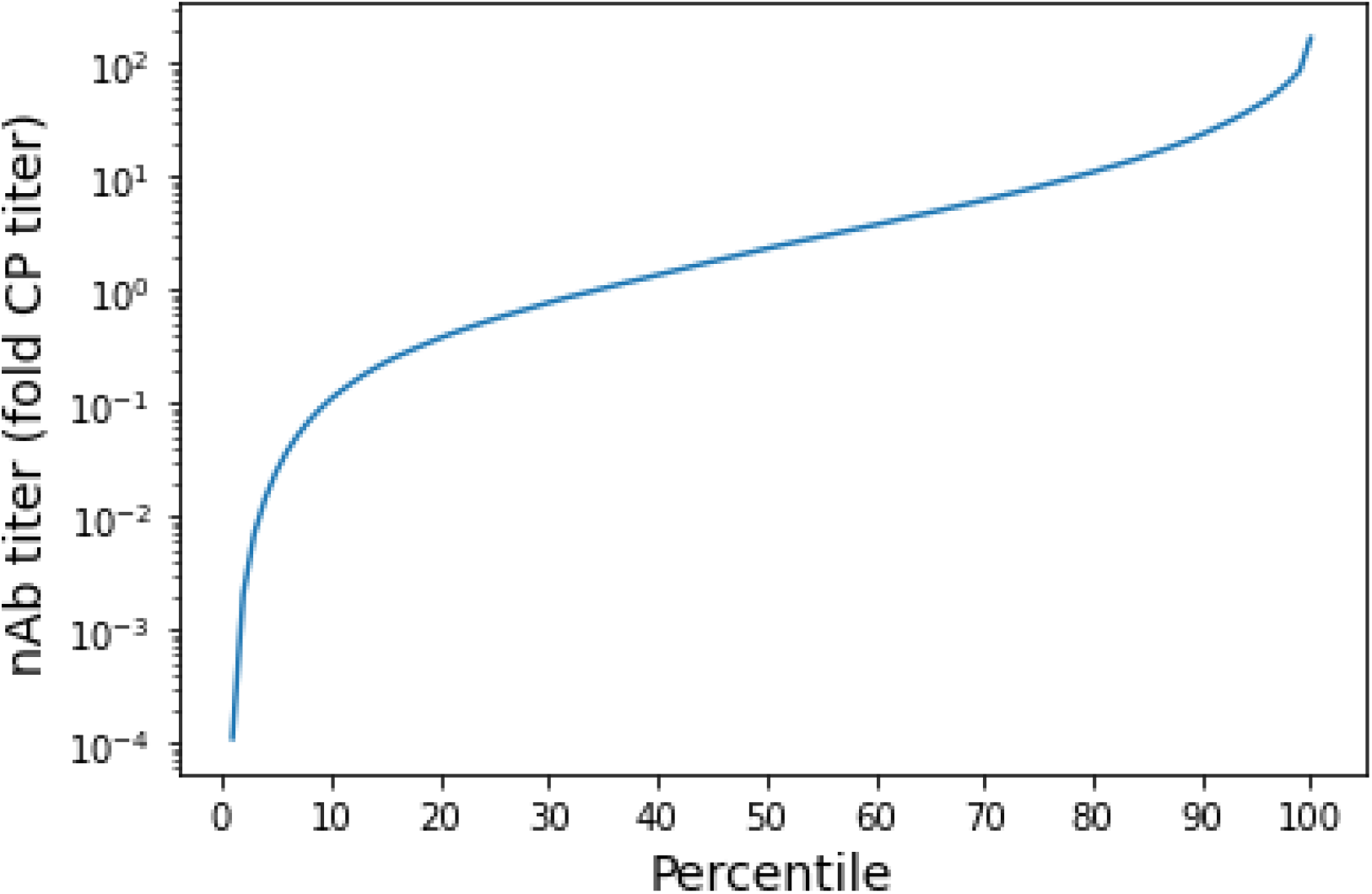
Percentile distribution of nAb titers in the population under endemic SARS-CoV-2 transmission. Titers are normalized against peak post-infection (CP) titer.

### Achieving reduction in SARS-CoV-2 infection frequency requires greater-than-proportional reduction in contact rate

Figure 5 demonstrates the nonlinear relationship between contact rate and reinfection frequency under endemic conditions: a given percent reduction in contact rate achieves a lesser percent reduction in infection frequency. For unvaccinated individuals with the median contact rate (i.e. an average person), an approximately five-fold reduction in contact rate is required to achieve a 50% reduction in infection frequency (from 2.1 times yearly to once yearly, Figure 5A). For vaccinated individuals, infection frequency is lower than the unvaccinated across all contact rates (Figure 5B, p < 10^−10^). However, a nearly 10-fold reduction in contact rate relative to the median is required to achieve a 50% reduction in infection frequency for the vaccinated (from 1.3 times yearly to 0.65 times yearly). Regardless of vaccination status, individuals with contact rates equal to 50% of the median experience infection frequencies only marginally less than individuals with contact rates equal to the median.

**Figure 5. :**
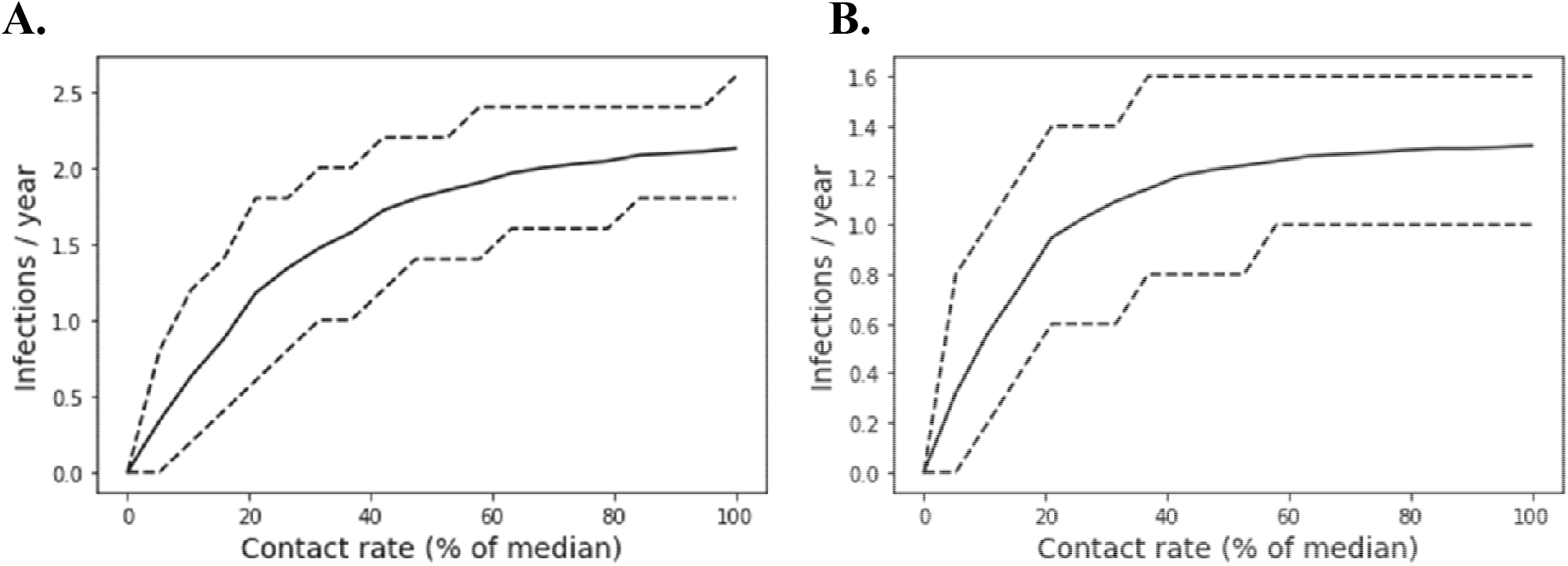
Frequency of infection (infections per year) is nonlinearly associated with individual contact rates. Infection frequencies among **A**. unvaccinated individuals and **B**. vaccinated individuals. Only individuals restricting their contact rates to less than 25% of the median achieve reductions in infection frequency. Solid lines represent the mean outcome while dashed lines delineate the 90% population interval.

Under endemic conditions, the relationship between contact rate and frequency of infection is nonlinear because of the maintenance of infection-induced immunity in the population. While reducing one’s contact rate prevents infection, immunity also prevents infection for a period of time. As a result, for individuals infected at any frequency, post-infection immunity dilutes some of the impact of reductions in contact rate. Thus, in a population with a high frequency of infection, the reduction in infection frequency achieved by a reduction in contact rate is smaller than the reduction in contact rate.

### Substantial steady-state burden of long COVID predicted among vaccinated and unvaccinated

SARS-CoV-2 infection imposes a risk of long COVID. In this analysis, we used data on long COVID risk after infection published by the Minneapolis Fed ^58^ to estimate the prevalence of long COVID of varying severity under endemic SARS-CoV-2 spread. Optimistically, we assumed a constant rate of recovery for long COVID based on the observed frequency of recovery after three months. Under these assumptions, we found that at steady-state, approximately 25% of unvaccinated individuals with the median contact rate have long COVID at any given time, while 12% of vaccinated individuals have long COVID (Figure 6). Greater than 5% of unvaccinated individuals and greater than 2% of vaccinated individuals with the median contact rate are predicted to experience long COVID so severe that it impacts their employment. While a minority of these individuals are expected to be fully unemployed, the study’s reported average impact was a 50% reduction in hours ^58^. Similar to the risk of infection, the risk of long COVID declines with reduced contact rates relative to the mean.

**Figure 6.**
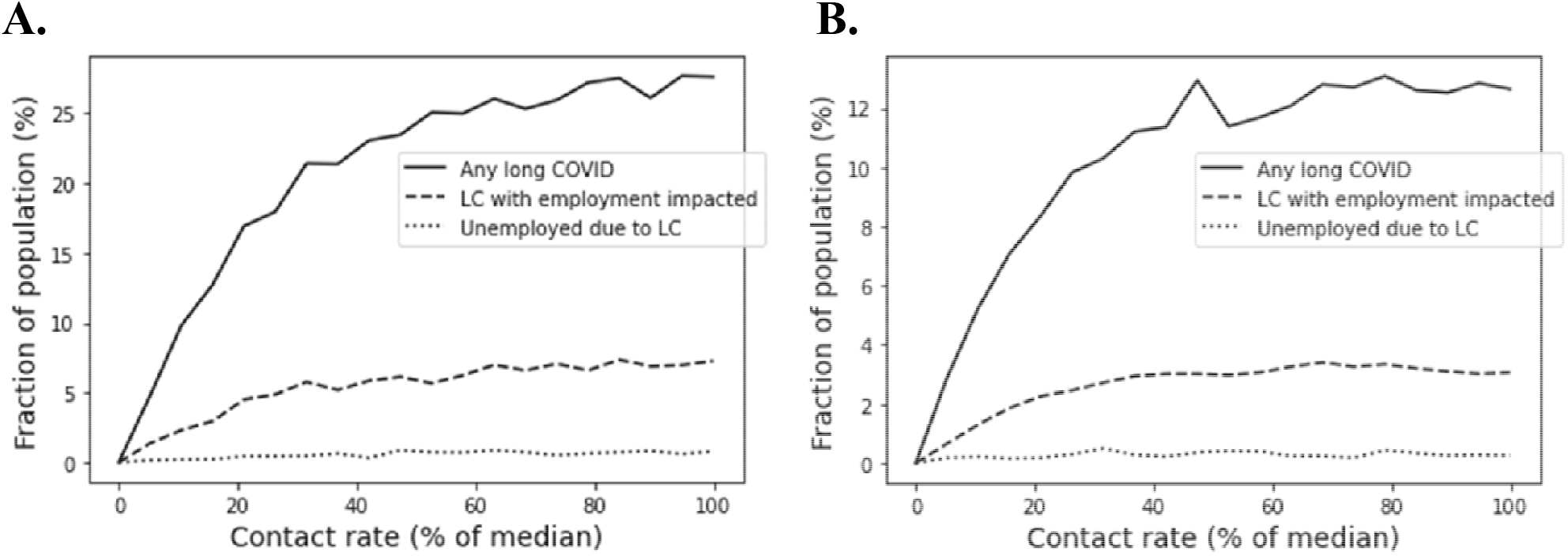
An individual’s long COVID risk decreases nonlinearly with reductions in contact rate. Fraction of **A**. Unvaccinated individuals and **B**. vaccinated individuals experiencing long COVID of varying severity at any given time as a function of individual contact rate.

## Discussion

At the current juncture of the ongoing COVID-19 pandemic, governments worldwide have mostly removed public health mitigation measures, placing the onus on individuals to protect themselves. In this work, we have examined the extent of contact reduction required to reduce frequency of infection using an agent-based model of an “endemic” SARS-CoV-2 scenario with repeated reinfections. We employed estimates of heterogeneity in immune protection derived from our population pharmacokinetic model to examine the protection afforded to individuals by their prior infections, with and without reductions in contact rate. As our understanding of the link between COVID-19 disease and downstream outcomes is still evolving, the study here relies on a number of simplifying assumptions. (For an in-depth discussion of the assumptions and the limitations of the study, see Supplementary Methods).

Our modeling indicates that the endemic steady-state will result in a large infection burden for both the vaccinated and unvaccinated population, with reinfection frequencies ranging from slightly less than once per year to more than twice yearly. This burden of disease has significant economic consequences, both at an individual and population level.

At an individual level, people who are vaccinated and not taking measures to reduce their contact rate can expect to spend an average of 6 days a year acutely sick with COVID-19 (1.4 infections/yr times 4.4 days average duration ^59^, and also incur a 12% risk of long COVID (symptoms lasting more than 3 months). Using the US as an example (where the average number of paid sick days is 8 ^60^, 27% of employees lack health insurance ^61^, 60% or more of employees lack short-term or long-term disability insurance ^61^ and average savings range from $3240 (under 35) to $6400 (55-64)) per person, many working individuals may find “living with COVID” to be beyond their means within a short period of time.

At a population level as well, the endemic steady-state will bring with it a substantial economic cost. Our modeling suggests that between 12 and 25% of the population (depending on up-to-date vaccination status) - 32 to 66 million adults-will have long COVID at any given time. Even if only a portion of those impacted are forced to leave their jobs or reduce working hours, the aggregate effect on labor supply, productivity, and income could be substantial. Already, various authors have estimated that the equivalent of somewhere between 500,000 and 4 million full-time workers have exited the labor force on account of post-acute COVID symptoms ^47,62,63^, with an additional group likely reducing its labor supply as well due to fears of contracting the virus on the job ^64^. The chair of the U.S. Federal Reserve recently posited that, insofar as long COVID can explain a portion of ongoing labor market tightness, it could also be one factor behind an increased rate of inflation ^65^.

We can expect wide variation in nAb levels within the population at the endemic steady-state due to heterogeneity in nAb persistence and contact rate. In keeping with this, there is substantial heterogeneity in the number of infections per year, in both the vaccinated and unvaccinated populations. Our results suggest that individuals seeking to limit their risk of infection have limited opportunities to “opt out”. Large reductions in contact rate are required to significantly reduce infection risk and the risk of long COVID.

These results have a number of practical implications for public health, as well as for individuals seeking to reduce their risk of infection or negative outcomes with COVID-19. At an individual level, large reductions in contact rate are necessary to reduce risk of infection. This is consistent with studies suggesting that the use of a high-quality (N95/FFP2+) mask is associated with a reduced risk of SARS-CoV-2 infection ^66–71^. On the other hand, partial measures may not be sufficient to reduce infection frequency despite reducing an individual’s contact rate. This is consistent with the observation that using a low-quality (e.g. surgical) mask ^72,73^ or masking intermittently ^74^. As a practical matter, individuals seeking to shield have a limited range of interventions available at this point to reduce risk of infection (vaccine boosters, one-way masking and social isolation), and many individuals are forced into high-contact settings as a result of work, school, medical need, or family commitments. As a result, the reduction in contact rate required to limit the risk of infection under unmitigated SARS-CoV-2 transmission is likely to be unachievable for many individuals. This highlights the limitations of a public health “individual responsibility” model focused on shielding as a practical strategy for limiting the health costs of COVID-19.

At a public health level, our work demonstrates that “personal choice” for COVID-19 outcomes under endemic conditions will require a societal commitment that is different from the situation in most countries at present. In this context, a minimal suite of public health interventions should be implemented to allow individuals to achieve the reduction in contact rate required to opt out of frequent reinfections and long COVID. These measures, for example, could include investing in improved air quality in public spaces and providing access to high-sensitivity rapid tests to limit in-household transmission. In addition, from the standpoint of disability protections, public health authorities should take steps to create conditions that permit the clinically vulnerable to reduce their rate of contact, such as implementing “masked-only” hours at essential venues (grocery stores, government buildings, pharmacies, medical facilities, public transit), encouraging low contact (virtual, delivery, outdoor, or curbside) options for accessing goods and services, and providing and protecting remote work and schooling options. Rampant nosocomial infection should be curtailed through masking in hospitals, surveillance testing, improvements in air quality, and separation of patients ^75–77^. Supporting individuals who seek to reduce their contact rate provides a low-cost and low-effort public health intervention.

As a further point for public health, it bears mentioning that individual measures may be effective in reducing an individual’s contact rate without impacting their frequency of infection. This suggests that measures be considered in terms of a suite of interventions (for example masking, indoor air quality and testing), rather than individually. Our findings linking reduction of contact rate to reduction in risk of infection also provide an explanation for the seeming paradox of the use of face masks-while masks have been compellingly demonstrated to reduce the efficiency of onward viral transmission using a physical sciences framework ^14,78,79^, observational studies have confirmed this impact in some cases ^66–71^, but not others ^80,81^. Notably, surgical masks have a minimal impact on reducing risk of infection ^72,73^, as does using a high-quality mask inconsistently (or sometimes) ^74^, exactly as would be expected based on the results presented here.

In the absence of concerted public health actions to provide options for individuals looking to reduce their risk of infection, shielding as a personal responsibility is challenging to achieve. Public health organizations have advocated for oxymoronic “individual public health measures”, but these represent an inadequate solution to the ongoing COVID-19 public health crisis. Permitting the unrestrained spread of SARS-CoV-2 in the population will inflict a heavy burden of infection and long COVID on society as a whole, which will be challenging if not impossible for individuals to opt out of in the long run.

## Methods

### Population mixed effects model fit for neutralization potency over time

To examine the kinetics of serum nAb titers after SARS-CoV-2 infection, we applied a two-stage model structure to the nAb titer dataset published by Wang et al ^54^. The two stages of the model are the production phase, in which antibody titers are both produced and eliminated, and the memory phase, where antibodies are only eliminated. The transition from the production phase to the elimination phase occurs at time T. We compared first-order and zero-order production models, and the zero-order production model was selected for neutralization potency based on low AICs and good parameter estimation with low standard errors (Table S2). The following equations describe the zero-order and first-order models:

Zero-order:

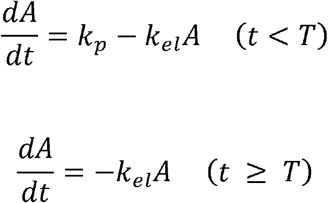

First-order:

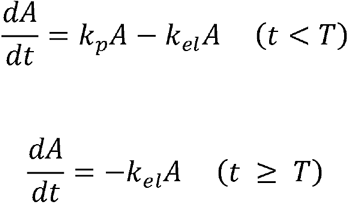

Where A is the antibody titer, k_p_ is the rate of antibody production, k_el_ is the rate of antibody elimination, and T is the duration of antibody production. The dataset contains nAb titers for 30 patients who recovered from COVID-19 and were discharged from Yongchuan Hospital of Chongqing Medical University. Age and gender were statistically analyzed as covariates in

Monolix. A goodness-of-fit analysis confirmed model specification and fit to the dataset (Figure S3).

### Half-life and peak titer calculation

Antibody half-life for individuals was calculated from individual decay rate according to the relation t_1/2_ = ln(2)/k_el_. The peak antibody titer for individuals can be calculated directly in the structural model by integrating the increase of antibody concentration over the time of antibody production. The distributions of individual half-lives and peak titers were visualized as probability density functions (PDFs).

### Population protection from symptomatic and severe COVID-19

To determine population heterogeneity in protection from symptomatic and severe COVID-19 over time after infection, we used the established relationship between neutralizing antibody titer and extent of protection ^55^. First, we used Monolix to simulate a population of 200 individuals drawn from the model-fitted parameter distributions. We simulated the neutralizing antibody titers of these individuals over two years after infection. We normalized the titers by peak convalescent plasma level – the average titer in the population at 34 days – in accordance with Khoury et al ^55^.

Normalized neutralizing antibody titers were transformed into levels of protection as follows:

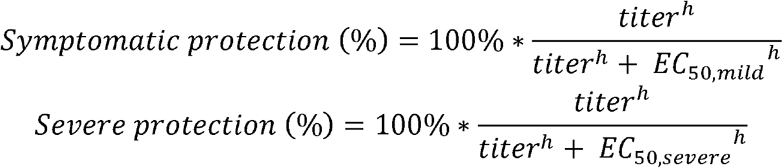

Where titer is the neutralizing antibody titer, h is the Hill coefficient, and EC_50_s for mild and severe disease are sourced from Khoury et al ^55^. For the entire time series, we sorted the protection levels by percentile and visualized the 10^th^, 25^th^, 50^th^, 75^th^, and 90^th^ percentiles. The population mean was calculated by averaging over all 100 percentiles.

### Agent-based simulation of SARS-CoV-2 dynamics

Under endemic conditions, individuals in the population will become reinfected with SARS-CoV-2 as their neutralizing antibody titers wane. To simulate the impact of heterogeneous natural immunity on long-term SARS-CoV-2 infection dynamics and severity, we developed a simplified agent-based epidemiological model that accounts for interindividual heterogeneity in the rate of antibody waning and in exposure risk (contact behavior). This model tracks the nAb titers of simulated individuals with fixed contact rates, neutralizing antibody decay rates, and vaccination statuses. The individuals’ contact rate and antibody decay rate are parameterized by a random draw of the contact distribution ^82^ and the neutralizing antibody half-life distribution derived from the population PK model fit. Contact rates are treated as relative rather than absolute – the model’s intrinsic reproductive number (R_0_) multiplied by the normalized individual contact rate determines the individual’s absolute rate of exposure.

For each individual, the cumulative number of infections, long COVID status, and neutralizing antibody titer are tracked over time. The neutralizing antibody titer of each individual wanes according to the individual’s decay rate. For each individual, titer is updated at

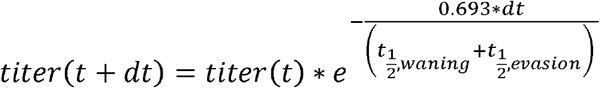

Where t_1/2, waning_ is the individual’s neutralizing antibody half-life and t_1/2, evasion_ is the half-life of nAb potency due to immune evasion.

Additionally, neutralizing antibody titers are boosted by a fixed multiple when the individual is successfully infected after an exposure or after vaccination. All individuals in the population are exposed at a rate proportional to their contact rate. This is implemented as a random draw on the population, with each individual’s likelihood of being drawn proportional to their contact rate. Upon exposure, an individual’s risk of infection is calculated based on their neutralizing antibody titer. The risk of infection upon exposure is:

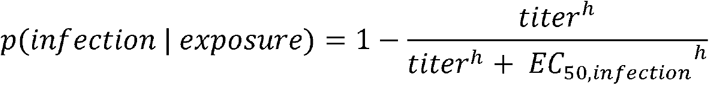

Where titer is the individual’s neutralizing antibody titer normalized to peak convalescent level, h is the Hill coefficient, and EC_50,infection_ is the neutralizing antibody titer required for 50% protection from infection. EC_50,infection_ was assumed to be 35% of the peak convalescent plasma level because we found this to be approximately consistent with clinical data suggesting median protection from reinfection is 83% over 5 months after infection ^83,84^. The exposure results in successful infection if a random value between 0 and 1 is less than p(infection | exposure). Then, the individual’s antibody titer is boosted by a fixed multiple ^55^.

Additionally, the individual may or may not develop long COVID upon successful infection. We based the risk of long COVID over time on the data published by the Minnesota Fed ^58^. This study estimates that 24.1% of COVID-19 cases result in long COVID lasting at least 3 months from infection. Optimistically, we assumed that long COVID only results from symptomatic infections, while approximately 67% of SARS-CoV-2 infections are symptomatic ^85^. Additionally, the study indicates that approximately 50% of long COVID patients had recovered, with a mean duration of six months (three additional months after the three-month minimum duration for long COVID ^58^). We used this point estimate to determine the risk of long COVID symptoms over time since infection, assuming an equal daily probability of recovery.

Thus, we estimated a daily probability of recovery of 0.38%, beginning at the three-month point. The risk of long COVID is reduced by 41% in the vaccinated subpopulation ^86^. If the individual acquires long COVID, the model determines the severity based on the distribution described in the study (25.9% of long COVID patients experience employment impacts, 10% of whom are unemployed ^58^. In the agent-based simulation, we tracked the severity of each case of long COVID, with the possibility of severity increasing if a pre-existing case has not resolved before the onset of another.

We assumed that all infections are of the same duration, and the number of active infections is tracked using a counter. The counter is increased by one for each successful infection, while recovery events decrease the counter by one. Each active infection has a 10% likelihood of recovery per day, based on an average 10-day infection duration ^87^. The rate of exposures is determined by the number of active infections multiplied by the R_0_ (the force of infection, or the expected number of secondary infections in the absence of pre-existing immunity). This determines how many exposures occur at each timestep, with exposures either succeeding or failing to induce an infection as described previously. As such, we do not account for overdispersion (variability in infectivity of individuals), and we assume that the population is well-mixed (any individual can infect any other individual, with no network effects).

Vaccination is modeled similarly to infection, with neutralizing antibody titers increased by a fixed multiple upon vaccination. Vaccine antibodies are assumed to decay at the same rate as antibodies from natural infection, which is somewhat optimistic ^88^. Each member of the simulated population is randomly designated as “vaccinated” or “unvaccinated” in a proportion consistent with the specified prevalence of vaccination in the population. All vaccinated individuals are boosted (revaccinated) on a fixed interval, which is tracked by an individual counter that numbers the simulation days since last vaccination. When the counter reaches the revaccination interval, the individual’s antibody titers are multiplied by the vaccination multiple and the counter is reset to zero.

Model parameters are summarized in Table 2.

**Table 2.** Agent-based model parameters.

| Parameter | Value | Source |
| --- | --- | --- |
| h | 1.018 | 55 |
| EC<sub>50, mild</sub> (CP titer*) | 0.22 | 55 |
| EC<sub>50, infection</sub> (CP titer*) | 0.35 |  |
| CP titer (ID50) | 643.2 | 54 |
| R0 (individuals) | 8.2 | 89,90 |
| Immune evasion half-life (days) | 73 | 91 |
| Infection duration (days) | 10 | 87 |
| Increase in titer on reinfection (fold) | 14.4 | 92 |
| Increase in titer on revaccination (fold) | 10 | 93–95 |
| Vaccine reduction in long COVID risk (%) | 41 | 86 |
| Revaccination interval (days) | 365 | 96 |
| Fraction boosted (%) | 50 | 56 |
| Long COVID risk upon infection (%) | 24.1 | 58 |
| Daily long COVID recovery probability (%) | 0.38 | 58 |
| Simulation duration (years) | 3 |  |
| Simulated population size | 100,000 |  |

\*Expressed as a fraction of peak neutralizing antibody titer in COVID-19 convalescents.

### Statistical significance

To address whether differences in outcome between groups with different individual parameters were statistically significant, we evaluated p-values for selected comparisons described in the Results sections and provided 90% population intervals in plots depicting the relationship between individual properties and outcomes. To calculate p-values, we used Welch’s t-test, which is an independent two-sample test that does not assume equal variance for the two samples. We did not perform any correction for multiple comparisons. To display the 90% population intervals, we sorted the range of COVID-19 outcomes in the population or subpopulation of each simulation run from greatest to least. Then, we binned the resultant distribution into 100 percentiles. We displayed the 5th and 95th percentiles, corresponding to the 90% population interval. This provides a visual means of assessing statistical significance.

## Data Availability

All data produced in the present study are available upon reasonable request to the authors

## Notes

### Competing Interest Statement

MS, LY, and AC are employees of Fractal Therapeutics, Inc. Fractal Therapeutics has no business interest in the topic of the article.

### Funding Statement

This study did not receive any funding

### Author Declarations

The study used ONLY openly available human data that were originally located at https://doi.org/10.1093/cid/ciaa1143

