## Supplementary Materials for "Shielding under endemic SARS-CoV-2 conditions is easier said than done: a model-based analysis"

**Affiliations:**

^3^ Independent Researcher

^4^ Stanford University School of Medicine, Stanford, CA, USA

^5^ Monash University, Melbourne, VIC, AUS

^6^ Halozyme Therapeutics, San Diego, CA, USA

^7^ Harvard Medical School, Boston, MA, USA

^8^ Boston Children’s Hospital, Boston, MA, USA

^9^ Sage Therapeutics, Cambridge, MA, USA

^10^ Boston University School of Public Health, Boston, MA, USA

* Corresponding author

 (A.C.)

**Abstract:**

As the COVID-19 pandemic continues unabated, many governments and public-health bodies worldwide have ceased to implement concerted measures for limiting viral spread, placing the onus instead on the individual. In this paper, we examine the feasibility of this proposition using an agent-based model to simulate the impact of individual shielding behaviors on reinfection frequency. We derive estimates of heterogeneity in immune protection from a population pharmacokinetic (pop PK) model of antibody kinetics following infection and variation in contact rate based on published estimates. Our results suggest that individuals seeking to opt out of adverse outcomes upon SARS-CoV-2 infection will find it challenging to do so, as large reductions in contact rate are required to reduce the risk of infection. Our findings suggest the importance of a multilayered strategy for those seeking to reduce the risk of infection. This work also suggests the importance of public health interventions such as universal masking in essential venues and air quality standards to ensure individual freedom of choice regarding COVID-19.

**Supplementary Materials:**

**A. B.**

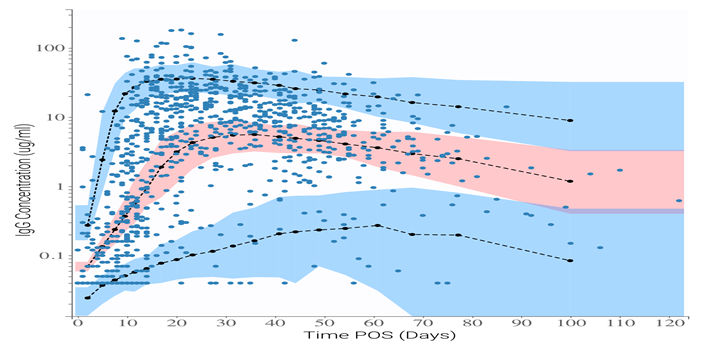

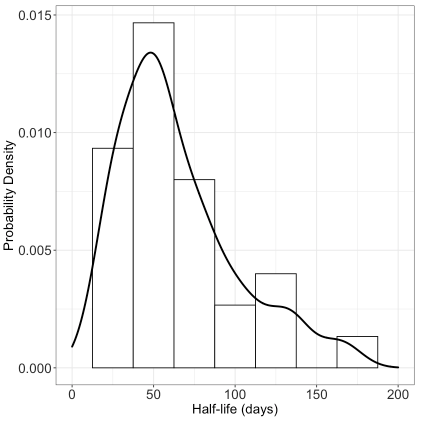

**Figure S1.** Mixed-effects model fit to IgG kinetics after SARS-CoV-2 infection. **A.** Visual predictive check for model agreement with fitted dataset. Shaded regions represent 90% prediction intervals for model certainty (pink area for the 50^th^ percentile, and blue areas for the 10^th^ and 90^th^ percentiles). Blue dots represent the fitted dataset, with dark blue lines representing the empirical 10^th^ and 90^th^ percentiles and red lines representing the empirical 50^th^ percentile. Distribution of **B**. IgG half-life probability density function.

**Table S2.** Parameter values for fitted IgG kinetics model with standard errors (SE) and relative standard error (RSE).

| **Parameter** | **Value** | **Units** | **Standard error** | **Relative standard error (%)** |
| --- | --- | --- | --- | --- |
| *Fixed effects (median)* | | | | |
| k_p, pop_ | 0.273 | IC50/days | 0.0367 | 13.4 |
| k_el, pop_ | 0.00998 | 1/days | 0.00182 | 18.3 |
| T_in, pop_ | 11 | days | 1.64 | 14.9 |
| *Standard deviation of the random effects* | | | | |
| ω_k_ | 0.602 | IC50/days | 0.0916 | 15.2 |
| ω_kel_ | 0.701 | 1/days | 0.174 | 24.8 |
| ω_Tin_ | 0.662 | days | 0.106 | 16 |
| *Correlations* | | | | |
| corr_k,Tin_ | -0.743 |  | 0.0986 | 13.3 |
| *Error model parameters* | | | | |
| b | 0.405 |  | 0.0323 | 7.98 |

where corr_k,Tin_ is the correlation between k_p_ and T_in_, and a is the coefficient of proportional error.

*Mixed effects model fit to IgG kinetics*

Based on a lower AIC, we found that the first-order production model best fitted the IgG kinetics data. No correlations between parameters or between parameters and covariates (age and gender) were found. The model is in good agreement with the data according to the visual predictive check (Figure S1A), and standard errors for parameter estimates are small, indicating appropriate model specification. Based on the model fit, the median IgG half-life is 64 days (24 days – 134 days 90% population interval, Figure S1B).

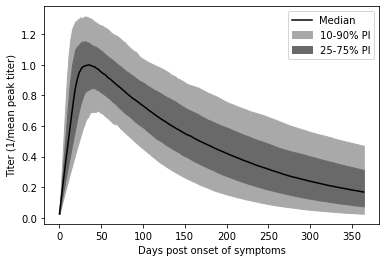

**Figure S2.** Normalized nAb titers post-onset of COVID-19 symptoms (POS) by percentile.

*After SARS-CoV-2 infection, neutralizing antibody titers wane across the population*

Figure S2 shows simulated antibody titers by percentile over time. Titers are normalized to the peak mean convalescent level, which is the mean titer at 34 days after onset of symptoms. A wide range of titers are observed at all timepoints, indicating significant variation in neutralizing antibody kinetics. In the 50^th^ percentile, titers wane by approximately 75% within one year of symptom onset. By this point, neutralizing antibody titers have waned by more than 10-fold in the 10^th^ percentile, while titers remain at about 60% of the mean peak convalescent level in the 90^th^ percentile. These results suggest wide variation in peak neutralizing antibody titers and waning kinetics.

**A.**

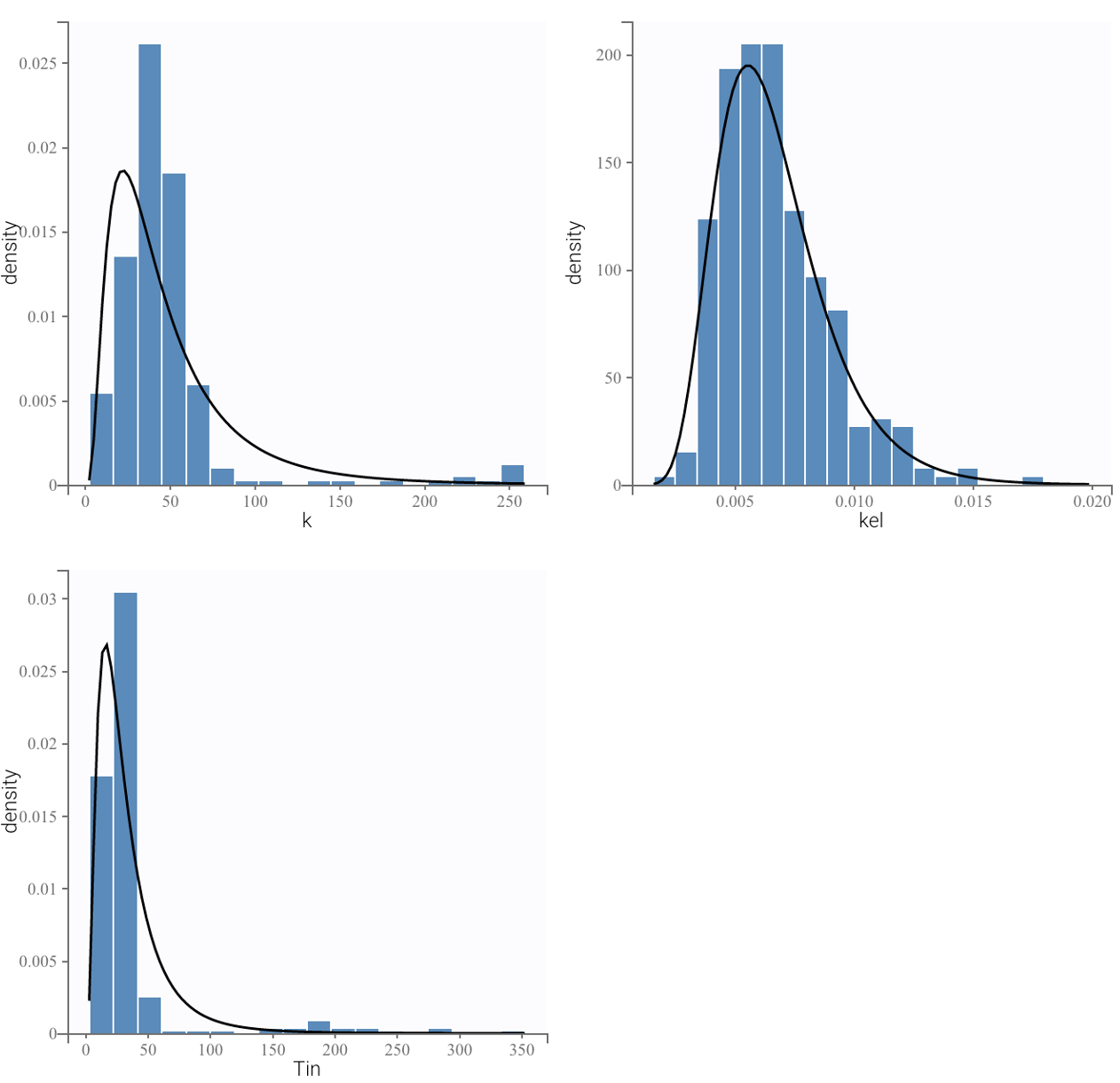

**B.**

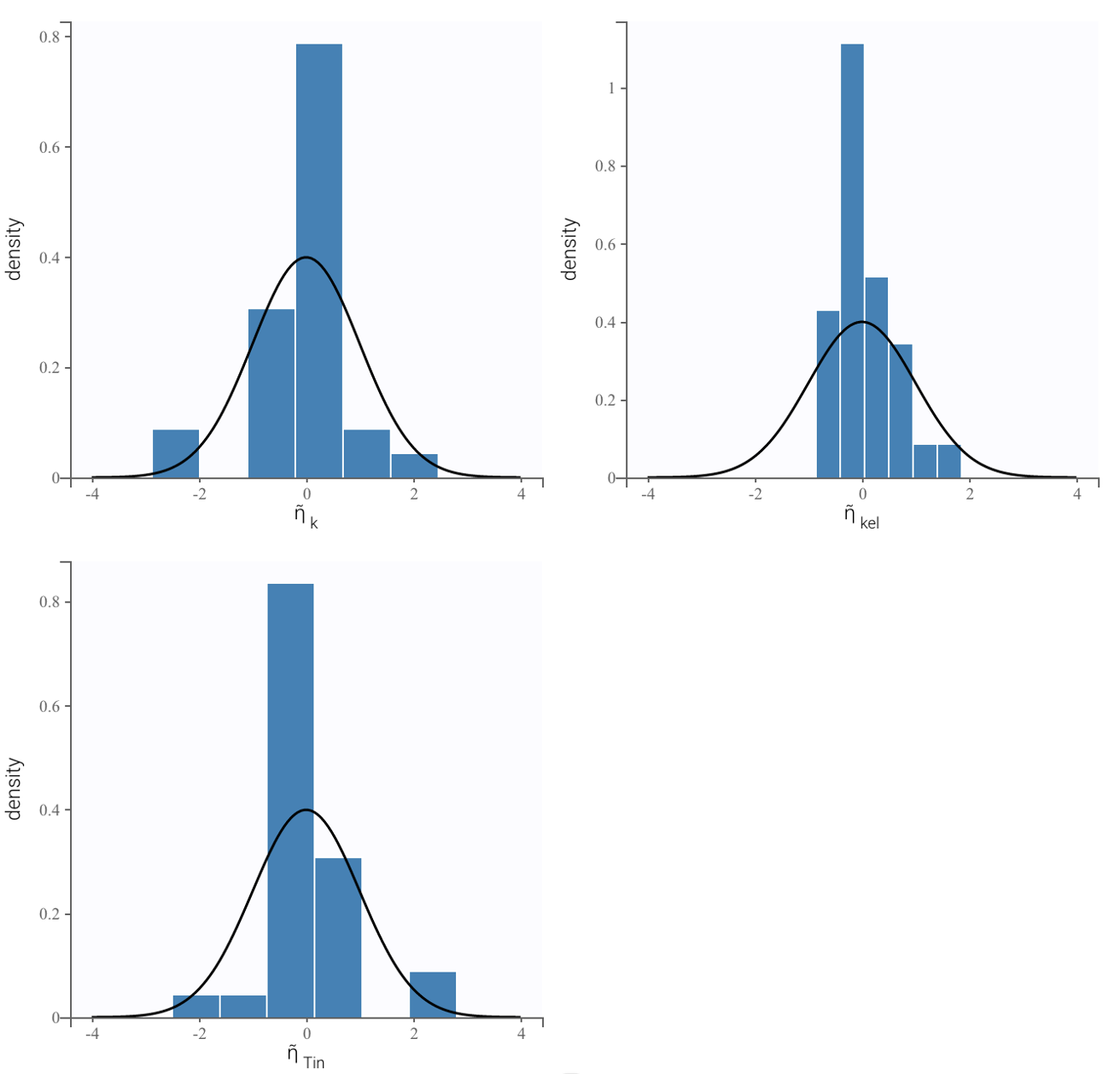

**C.**

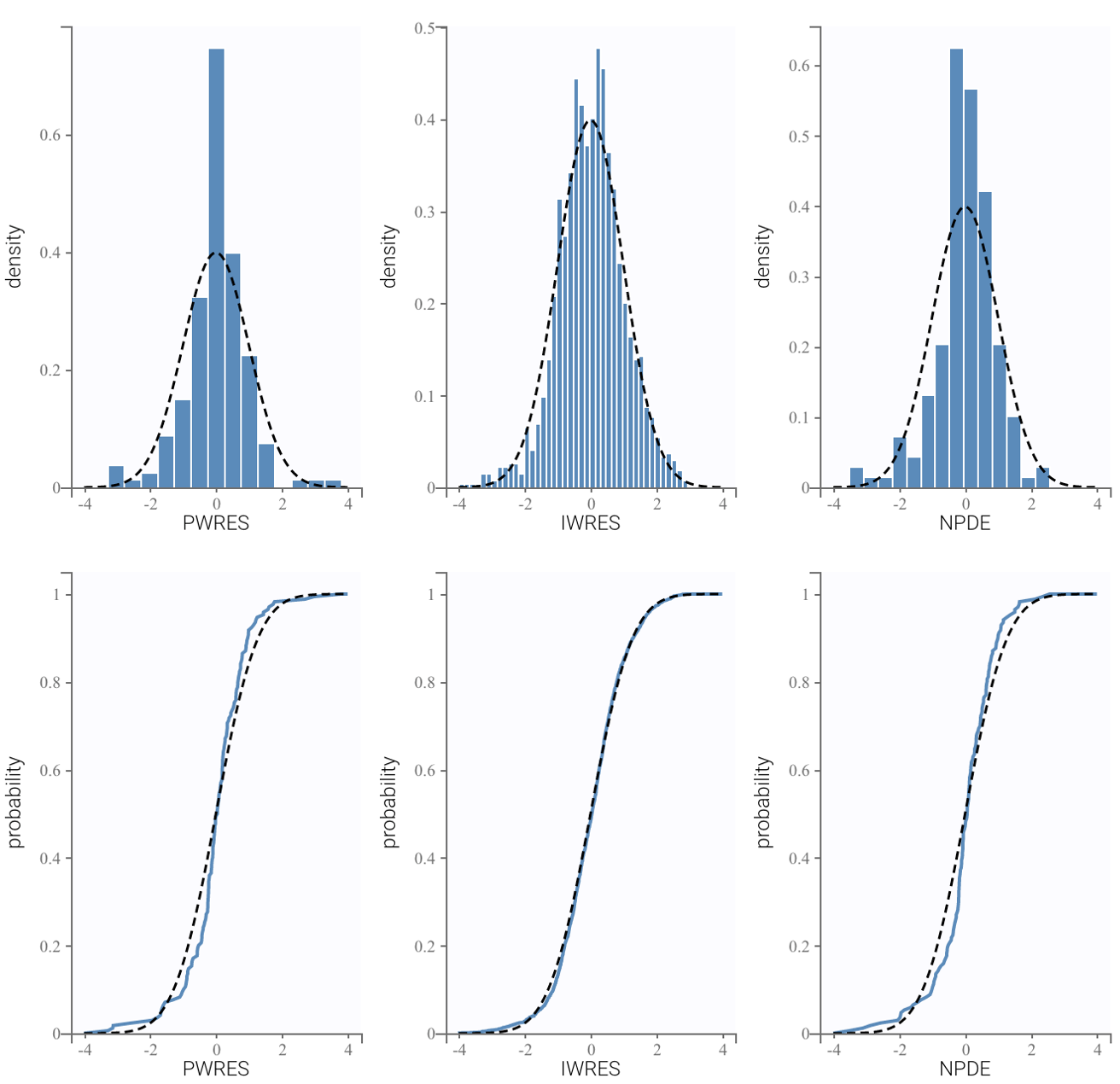

**D.**

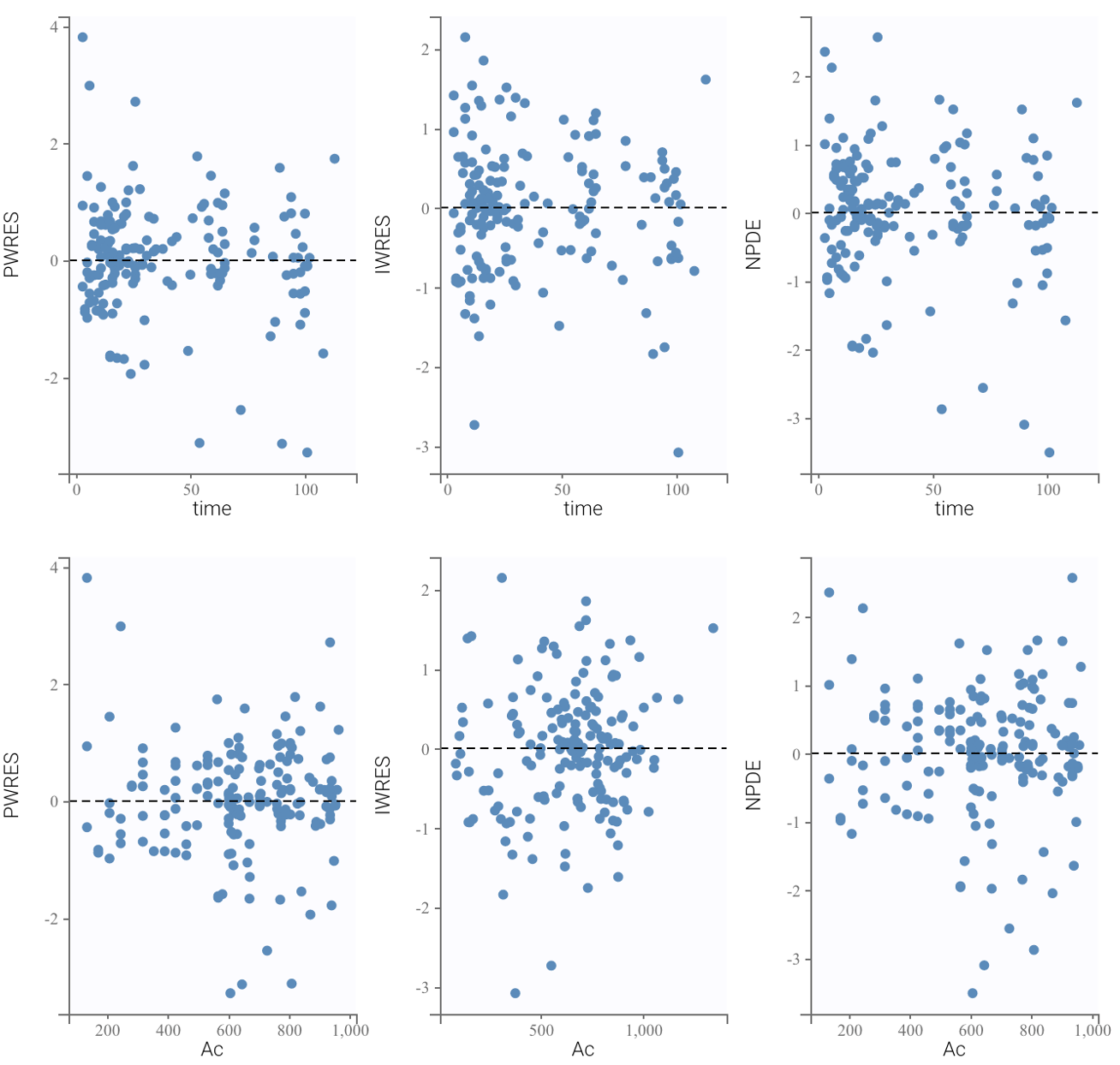

**E.**

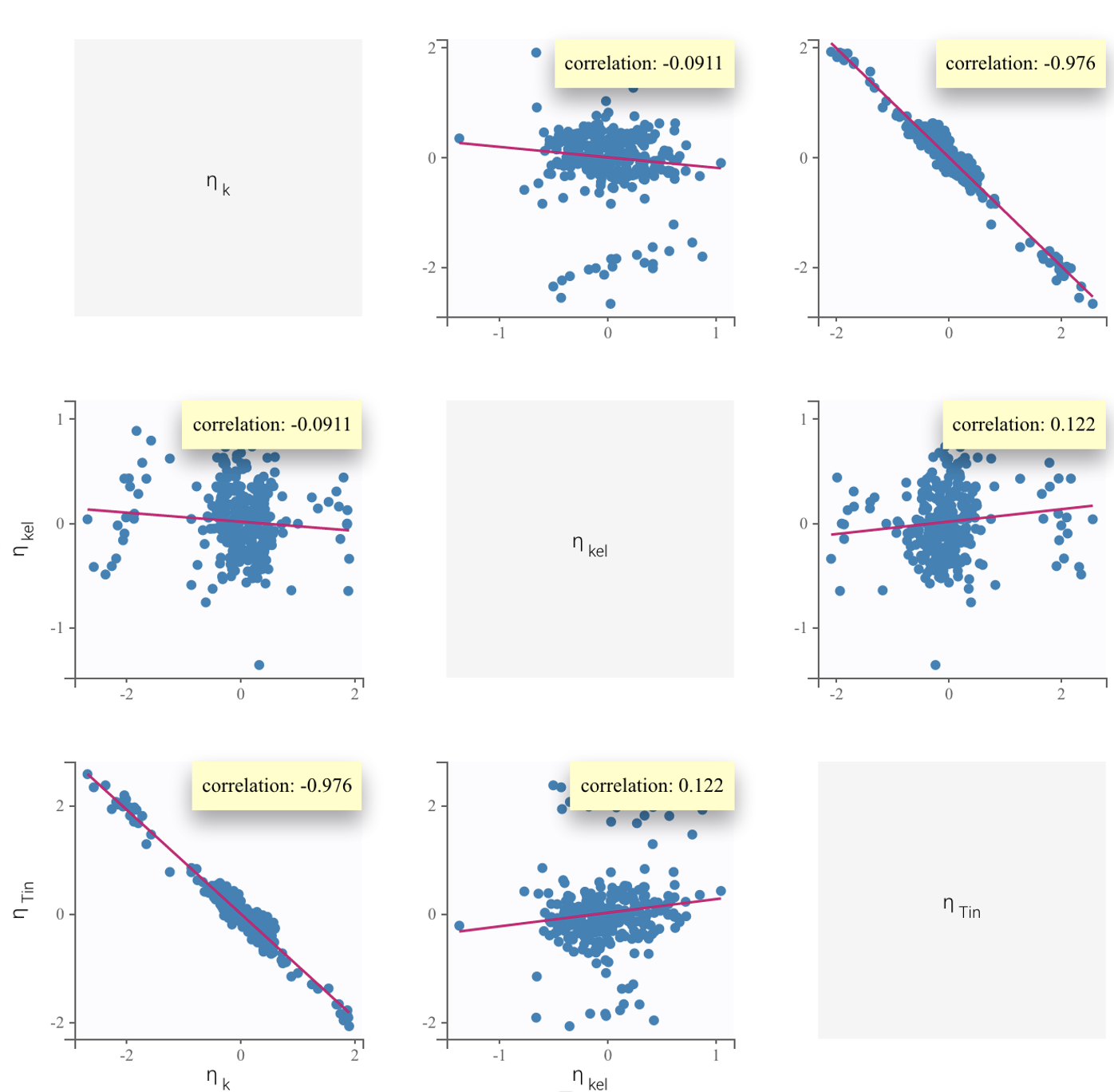

**Figure S3.** Goodness-of-fit assessment for post-COVID-19 nAb kinetics model. **A.** Probability distribution of individual parameters. The histogram plots are empirical distributions. The black line shows the theoretical distribution defined in the statistical model, which is a log-normal distribution. **B.** Probability distribution of standardized random effects. The histogram plots are empirical distributions. The theoretical distribution is shown as a dotted curve which is a normal distribution. **C.** Scatter plots of the residuals. These plots display the PWRES (population weighted residuals), the IWRES (individual weighted residuals), and the NPDEs (normalized prediction distribution errors) as scatter plots with respect to the time and the prediction. residuals should be randomly scattered around the horizontal zero-line, which means the selected proportional error model is true. **D.** Distribution of the residuals. Empirical and theoretical probability density functions (PDF) of the PWRES, IWRES and NPDE are on top. Empirical and theoretical cumulative distribution functions (CDF) are at the bottom. Normally distributed residuals indicate that the selected error model is true. **E.** Correlations plots between model kinetic parameters. Solid lines represent the mean outcome while dashed lines delineate the 90% population interval.

**Table S2.** AIC values for prospective model structures.

| **Object** | **Model** | **AIC** |
| --- | --- | --- |
| Nabs | 0-order production | 2271.82 |
| Nabs | 1st-order production | 2315.19 |
| IgG | 0-order production | 1275.52 |
| IgG | 1st-order production | 1253.82 |

**Table S3.** Half-lives of antibodies raised against viruses.

| **Virus against which antibodies are raised** | **Antibody half-life** | **Additional comment** |
| --- | --- | --- |
| ***SARS-CoV-2 and other viruses with relatively short antibody half-lives (days)*** | | |
| SARS-CoV-2 | nAb half-life = 109 days  IgG half-life = 64 days | Present analysis |
| SARS (2003) | nAb half-life = 6.4 weeks | Analysis was done using linear mixed model and SARS database ^1^ |
| Common cold coronavirus  (229E) |  | nAbs and specific IgG and IgA in sera were significantly raised in sera taken about 3 weeks after inoculation with 229E, but by 12 weeks the concentrations had declined considerably ^2^ |
| Influenza | Binding antibody half-life = 185 days ^3^ |  |
| Respiratory syncytial virus |  | Rapid decay of nAbs within a few months months^4,5^ to one year of infection facilitates reinfection^6^ |
| Dengue virus | Binding Ab half-life = 4 yrs ^7^ |  |
| ***Cohort 2: Viruses with relatively long antibody half-lives (years)*** | | |
| Yellow fever |  | nAbs to 17D YF (17D yellow fever) virus vaccine persist for 30 years ^8^ |
| Vaccinia  (Smallpox vaccine) |  | Serum antibody levels were exceptionally stable between 1 and 75 years after vaccination, thus making the determination of the half-life of antibody decay impossible ^9^ |
| VZV  (Varicella-zoster virus) | IgG functional half-life = 50 yrs | The long half-life is observed in the absence of boosting, hinting at the presence of plasma cells that are long-lived ^10^ |
| Measles | Estimated half-life = 3014 yrs ^11^ |  |
| Mumps | Estimated half-life = 542 yrs ^11^ |  |
| Rubella | Estimated half-life = 114 yrs ^11^ |  |

**Supplementary Methods:**

*Notes on assumptions and limitations of this study*

The model in this paper addresses multiple sources of variation in COVID-19 risk in the general population: heterogeneity in neutralizing antibody decay rate, contact rate, vaccination status, and stochastic risk of exposure. To achieve this, we have made two key assumptions in the context of a standard agent-based epidemiological model. Firstly, we assumed that the increase in titer upon vaccination or reinfection is a fixed multiple. In the model, this drives very high nAb titers in some individuals, which are generally consistent with titers observed among individuals with “hybrid immunity” ^12^. Secondly, we assumed that upon SARS-CoV-2 exposure, nAb titer alone determines risk of infection according to the model published by Khoury et al ^13^. Lastly, we assumed that vaccination reduces long COVID risk independently of nAb titers by a fixed 41% ^14^, although estimates vary ^15^.

Several factors impacting an individual’s burden of COVID-19 are not addressed in this work. We did not address the likely heterogeneity in peak immune response to reinfection or vaccination due to a lack of data regarding peak titers achieved after repeated infection and vaccination. We simplified immune evasion to a constant rate of antibody decay, and this rate may be optimistic. In a separate work, we have examined the implications of discrete jumps in immune evasion on public health outcomes (manuscript in progress). We have also not examined the impact of more frequent vaccine boosting schedules – this has been explored in other analyses (^16^, manuscript in progress). We used optimistic assumptions about boosters’ impact on nAb titer (a fixed 10-fold increase in titer every time, consistent with the 4th booster ^17^ and vaccine-induced nAb half-life (assumed to be the same as natural immunity, while vaccine titers wane more rapidly with more variability ^18,19^.

As a limitation, our work assumes that the viral inoculum does not influence risk of infection. However, some evidence suggests that a greater viral inoculum may increase risk of infection and severe disease ^20,21^. This may pose a greater risk to individuals with many contacts or who spend time in high-risk environments.

The agent-based model developed here assumes the simulated population is well-mixed. In this respect, the model is simplified compared to other agent-based models like Covasim, which includes transmission networks representing schools, workplaces, and other venues ^22^. Instead, our model narrowly addresses heterogeneity in contact rates, immunological parameters, and vaccination status and focuses on interindividual variation in immunity dynamics at steady-state. Our model, therefore, shares some limitations with compartmental SIR models ^23,24^. In particular, these models may overestimate the peak number of infectious individuals during the epidemic stage ^25^. However, we minimize the impact of these limitations by focusing the analysis on steady-state conditions.

Of note, we have used nAb titers here as a correlate of immune protection and neglected any potential contribution of other arms of the immune system, such as T-cells. Neutralizing antibody titers have been demonstrated to be a correlate of immune protection for SARS-CoV-2 ^13,26–31^, as they have for other viruses ^32^. A recent meta-analysis of nAb titers normalized to the mean convalescent titer (from the same study) showed a strong nonlinear relationship correlating with reported vaccinal protection across a range of different vaccines (Khoury et al., 2021).  Dose-response relationships were established both for protection against infection and severe COVID-19 outcomes. Neutralizing antibody titers correlate with waning vaccinal protection against infection (VEi) due to PK waning ^29,33^ or viral immune evasion ^34–38^ and with loss of vaccinal efficacy against severe disease (VEs) ^13^. The robustness of T-cell epitopes in the face of the newer immune-evading variants ^39,40^ has not translated into lasting vaccinal protection against infection or severe disease. T-cells are infected by SARS-CoV-2 ^41^, are functionally exhausted by severe COVID ^42,43^ and undergo frank apoptosis ^44,45^. Thus, a protective role for T-cell immunity for SARS-CoV-2 remains to be demonstrated.

*Notes on Long COVID*

Long COVID is a condition with complex, multifactorial pathogenesis (similar in this respect to other disease such as tuberculosis, asthma, heart disease, diabetes and cancer) ^46^. The term is used to describe a variety of post-acute sequelae of COVID. Reports indicate that a significant proportion of long COVID sufferers are impacted to the point of having difficulty with day-to-day functioning ^47–49^.

The evidence for hypotheses implicating an organic (physical) cause for long COVID is strong and growing: replicating virus has been found in autopsies of people who died with covid ^50^, viral persistence occurs even in mild cases of COVID-19 for up to two months ^51^, and viral persistence for over a year has been demonstrated in patients with long COVID ^52^. Consistent with the viral reservoir hypothesis, spike protein is found at elevated levels in the blood of ~60% of patients with long COVID ^53^.

In the study used to parameterize our model, the Minnesota Fed defines long COVID as "covid related symptoms or health complications that lasted at least 12 weeks" ^54^. Because the term ‘long COVID’ is used to mean different things at this point, estimates of the incidence of long COVID after infection vary widely ^47,55,56^. However, studies using similar definitions of long COVID to the Minnesota Fed study have yielded similar metrics. The Census Bureau’s Household Pulse Survey (HPS) found that around 8% of working-age Americans had long COVID (defined as symptoms lasting longer than 3 months) in June-July 2022 ^57^. The Federal Reserve Bank of Minneapolis study findings on long COVID (using the same definition of the condition) are in line with the HPS figures. (Specifically, at the time of the study, the CDC estimated that 70% of Americans had contracted covid at some point ^58^, yielding an estimate of 34 million Americans having had long COVID at some point). A number of other studies have yielded similar estimates of long COVID frequency. For example, the Brookings Institution report estimated that around 30 million Americans had had long COVID by January 2022). Similarly, United Kingdom’s National Health Services estimates that 3-11% of the UK population currently has long COVID (defined as “symptoms that persist 12 weeks beyond the acute phase of the ...COVID-19 infection ...headache, myalgia, fatigue, and loss of taste and smell. Parosmia can persist for months after initial infection alongside brain fog and memory loss" ^59^.

A number of studies have examined the employment impact of long COVID ^47,54,60^. However, because each of these studies has examined the problem through the lens of slightly different questions, the estimates are difficult to compare.

More generally, because the term ‘long COVID’ is used to describe a variety of different post-acute sequelae of COVID, estimates of the duration of long COVID are also varied. Disagreement is significant between studies, with some suggesting little decrease in long COVID symptoms over time since diagnosis ^61^ and others suggesting most long COVID sufferers recover within one year ^62^. A number of studies indicate that some long COVID patients do not see their symptoms resolve over longer periods of time. For example, one study found that >90% of respondents with long COVID required over 35 weeks for recovery ^47^. Another survey conducted by the Trades Union Congress (UK) found that 30% of their respondents had been experiencing long COVID symptoms for 12 months or more ^60^. However, surveys of long COVID may be confounded by response bias- response rate may be disproportionately high among those impacted most severely.

To minimize the impact of confounders as well as variable definitions of long COVID, we included concrete, employment-related outcomes as a measure of long COVID impact. We implemented long COVID incidence and employment impact estimates from a Minnesota Fed study based on longitudinal survey data ^54^. In a critique of this survey-based study, Salwati and Sheiner (2022) have argued that this survey data may overstate the reduction in working hours due to long COVID on account of non-representative sampling and/or lack of statistical power ^63^. However, their analysis relies on data from the Current Population Survey (rather than the HPS dataset that the Minneapolis Fed Study is based on), which does not explicitly ask about long COVID, and relies on a number of assumptions about what portion of reported disability cases are in fact COVID-related. Moreover, disability rates in the CPS have been shown to be lower than in other surveys ^64^.

In this paper, we have not considered acute post-COVID outcomes such as the increase in risk of heart attacks and strokes ^65^. These downsides stack on top of the long COVID risks we predict.
